# Genetic diversity and spatiotemporal distribution of SARS-CoV-2 alpha variant in India

**DOI:** 10.1101/2022.04.20.22274084

**Authors:** Jahnavi Parasar, Rudra Kumar Pandey, Yashvant Patel, Prajjval Pratap Singh, Anshika Srivastava, Rahul Mishra, Bhupendra Kumar, Niraj Rai, Vijay Nath Mishra, Pankaj Shrivastava, Prashanth Suravajhala, Gyaneshwer Chaubey

## Abstract

After the spill to humans, in the timeline of SARS-CoV-2, several positively selected variants have emerged. A phylogeographic study on these variants can reveal their spatial and temporal distribution. In December 2020, the alpha variant of the severe acute respiratory syndrome coronavirus (SARS-CoV-2), which has been designated as a variant of concern (VOC) by WHO, was discovered in the southeastern United Kingdom (UK). Slowly, it expanded across India, with a considerable number of cases, particularly in North India. The study focuses on determining the prevalence and expansion of the alpha variants in various parts of India. The genetic diversity estimation helped us understand various evolutionary forces that have shaped the spatial distribution of this variant during the peak. Overall, our study paves the way to understand the evolution and expansion of a virus variant.

## Introduction

The COVID-19 pandemic, caused by the SARS Coronavirus 2 (SARS CoV-2), has impacted the world, with India having the second-highest number of confirmed cases (*MOHFW-GoI*, n.d.). The first instances of COVID -19 in India were recorded in Kerala in January 2020 among three medical students who had returned from Wuhan, the pandemic’s first epicentre (Wu et al., 2020). SARS-CoV-2 is a single-stranded, positive-sense RNA (+ssRNA) virus in the Coronaviridae family that belongs to lineage B of the genus Beta-coronavirus. The genome size of SARS-CoV-2 is ∼29.9 kb, sharing ∼78% sequence homology with SARS-CoV (Qianqian Zhang et al., 2021). SARS-CoV-2 has been evolving at a rate consistent with the virus gaining two mutations per month in the global population since December 2019 (William T. Harvey et al., 2021). Mutation has given rise to different variants of SARS-CoV-2. Any virus strain becomes a variant of concern when it increases transmissibility and virulence or decreases its effectiveness towards vaccines (www.who.int, last accessed April 15, 2022.). The principal variants of concern are Alpha (B.1.1.7), Beta (B.1351), Delta (B.16172) and Gamma (P.1).

The Alpha VOC (lineage B.1.1.7) first appeared in the United Kingdom (Robert Challen et al., 2021) in South Africa, Beta VOC (lineage B.1.351), Gamma VOC (lineage B.1.1.28.1 or P.1) in Brazil (Finlay Campbell et al., 2021) and Delta VOC (lineage B.1.617.2) in India (Ewen Callaway, 2021). These VOCs were thought to be more resistant to the neutralizing activity of antibodies produced during natural infection or vaccination and their increased transmission rate (Gobeil et al., 2021). The viral Spike protein’s Receptor Binding Domain (RBD) contains most of the mutations observed in VOCs (S). Mutation N501Y, present in all except the Delta VOC, confers a higher affinity for the cellular ACE2 (Angiotensin-converting Enzyme 2) viral receptor and may be related to increased transmissibility of VOCs carrying this mutation (Rossana C.Jaspe et al., 2021). After being detected in the UK in no time, the Alpha variant became dominant in other countries. Its rapid success was due to its capacity to weaken our body’s first immune defense line, giving the mutant more time to reproduce. Alpha has 23 mutations, eight of which are in the spike protein, that distinguish it from other coronaviruses. N501Y, spike deletion 69-70del, and P681H are the three mutations that have the greatest biological impact. It was also found that there was an 80 fold increase in Orf9b which leads to an increase in Orf9b protein levels in this variant. Orf9b protein of virus antagonizes innate immune response by interacting with Tom 70, a mitochondrial import receptor required for Type I interferon production (Lucy et al., 2021). Beta and Delta differ from Alpha in terms of the production of Orf9b protein (Singh et al. 2020). They drive down levels of interferon but to do so, they do not overproduce Orf9 proteins.

According to the data obtained from GISAID (*GISAID - Initiative*, 2008), there is a variation in the number of alpha cases recorded in different parts of India. When the frequency of the Alpha variant was calculated across India, it was found that it is more common in various northern and central Indian states than in the southern states. In February and March 2020, several Indian states had the largest number of Alpha cases, after which India saw the second wave of the pandemic. Phylogenetic examination of samples from multiple Indian states revealed that it arose in the northern parts of the country from where it spread through the country. While affected regions vary in their value of genetic diversity, the relationship between genetic diversity and the number of cases provided a stance for these mutations in the conditions that prevailed in those regions, perhaps resulting in variable diversity in different regions.

## Material and Methods

GISAID, an open source for Covid 19 and influenza viruses, was used to download the genomic data of the alpha variant (*GISAID - Initiative*, 2008; www.gisaid.org, last accessed April 15, 2022). Multiple Alignment using Fast Fourier Transform (*MAFFT*, 2020) was used to align genome sequences to reference sequences. Aligned sequences were downloaded in Fasta format and were run on MEGAX (*MEGAX*, 2021) for the phylogenetic tree. Charts and Graphs were constructed by (*Google Sheets*, 2009). Frequency maps were generated by Datawrapper (2012), and haplotype diversity was derived from DnaSP (Julio Rozas et al., 2019)

1131 genomic sequences of the alpha variant were extracted on June 11/2021, with high coverage and aligned to the reference sequence (Wuhan/2019-EPI_ISL_402124). Each state, union territory and Wuhan sequences were labelled with distinct colours to be distinguished on a phylogenetic tree. Alpha variant samples were filtered from all the affected regions, and the frequency percentage was calculated. From December 2020 to May 2021, the frequency was determined separately for each month, and then distribution in different parts of India was evaluated. Its dominance in specific locations of India was discovered using the obtained frequency. Samples from each state and union territory were loaded on DnaSP to determine mutations in all impacted regions. The value of haplotype diversity was calculated using Fu and Li’s approach (Fu and Li 1993).

## Results and Discussion

Among the available sequences from India, Punjab had the highest number of samples for alpha variant with a frequency of 0.74, followed by Chandigarh (0.467) and Madhya Pradesh (0.296) (Supplementary Table 1). According to GISAID data, cases of alpha in India began to appear in a few states in October and November 2020, while the UK’s first sample was reported in September 2020 (A. Sarah Walker et al., 2021). In November 2021, samples of Alpha were discovered in the states of Delhi, Gujarat,, Kerala and Jammu & Kashmir, and it gradually spread to other regions of India.

Phylogenetic analysis revealed that Punjab and Delhi samples share a common ancestry at several nodes, indicating a link between the two regions and possibly the cradle of the virus spreading to other areas (Figure 1 and Supplementary Fig. 1). Samples from Sikkim, Chhattisgarh, Maharashtra, Gujarat, Jammu & Kashmir, and Uttarakhand shared a common ancestry with Delhi and Punjab (Fig. 1). For the temporal comparison, the month-by-month frequency was computed (Figure 2 and Supplementary Fig. 2). None of India’s zones had a high frequency of alpha in December and January, while Punjab saw a 0.354 frequency elevation in February. Along with Punjab, Madhya Pradesh has experienced an increase in cases. Except for the north-east states, nearly every region had the highest frequency in March. For example, in March, Punjab had a dramatic increase in frequency (0.6), followed by Chandigarh (0.294), Madhya Pradesh (0.214), and Jammu-Kashmir (0.16) (Figure 2).

**Figure 1.**
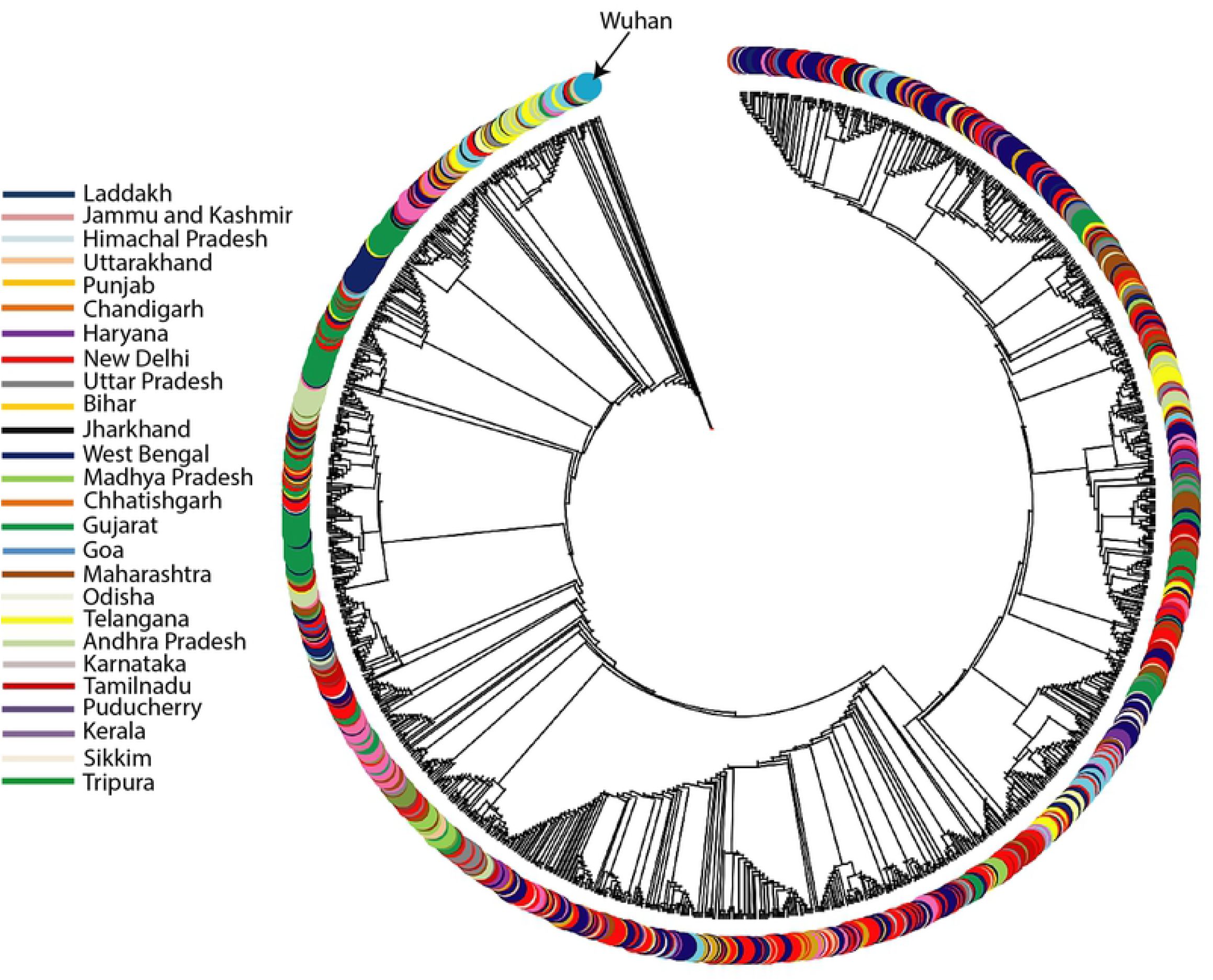
The Maximum likelihood (ML) tree of SARS-CoV-2 alpha (B1.1.7) variant showing the phylogenetic relationships of all the alpha variant samples reported in India. The samples have been segregated statewise.

**Figure 2.**
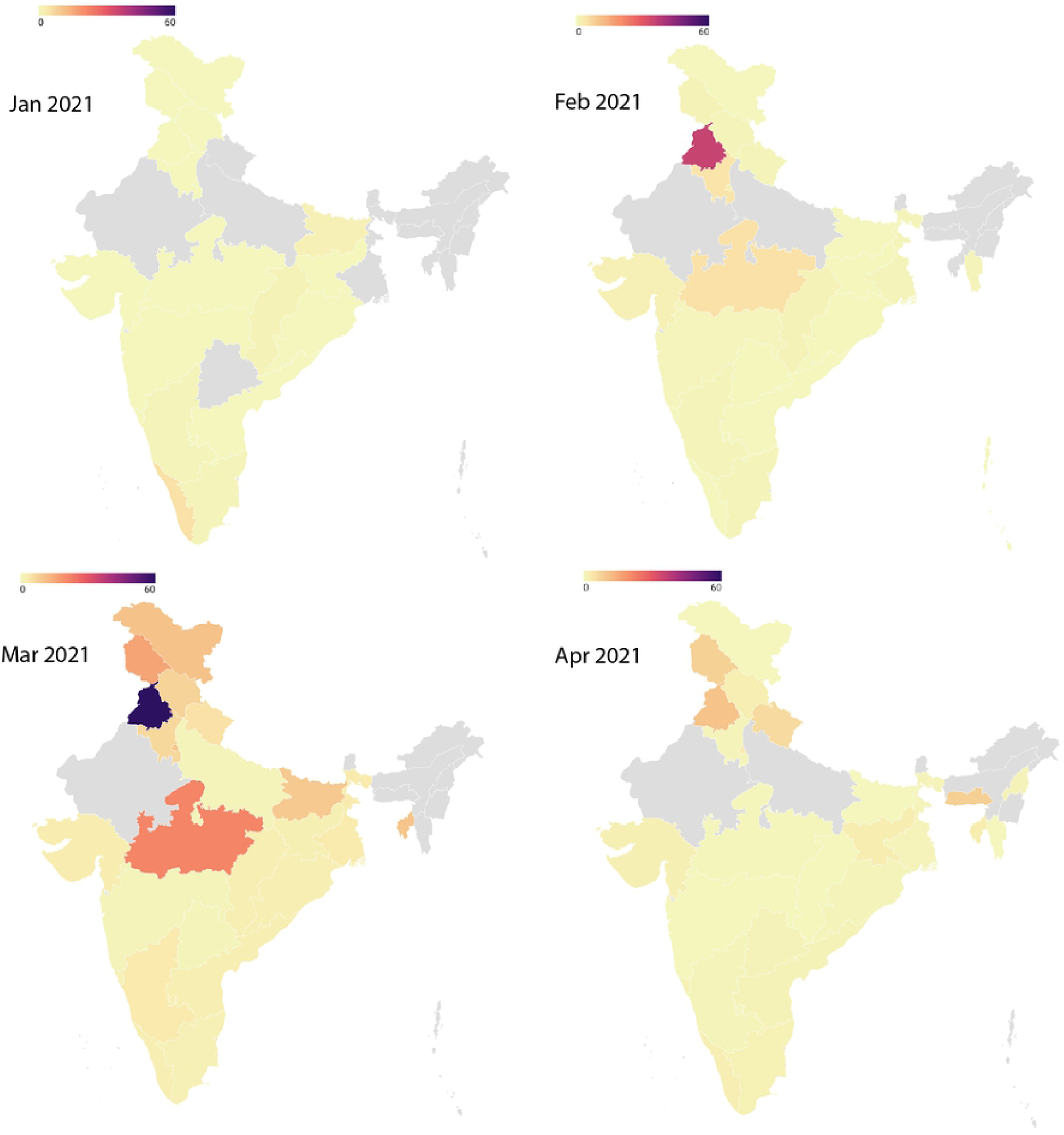
The geospatial frequency distribution of Alpha variant in India between January-April 2021.

In order to understand the diversity of this variant in different regions of India, we computed the haplotype diversity (Table 1). The diversity of this variant ranged from 0.798 to 0.997 in India. The highest haplotype diversity values were found in Chandigarh and Haryana (0.997), followed by Gujarat and Delhi, 0.995 (Table 1). This disparity in the diversity values range among various locations suggests variable viral dynamics in different regions (Table 1). Nearly high diversity in Chandigarh, Haryana, Gujarat and Delhi suggested either the multiple incoming or the prolonged presence of this variant and at least two of its lineages in these regions.

**Table 1.** Haplotype diversity of alpha variant (B1.1.7) in various regions of India. The values are arranged in descending order.

| States & Union Territories | Total Number of Samples | Haplotype Diversity |
| --- | --- | --- |
| Chandigarh | 92 | 0.997 |
| Haryana | 46 | 0.997 |
| Gujarat | 194 | 0.995 |
| New Delhi | 681 | 0.995 |
| Telangana | 74 | 0.993 |
| Andhra Pradesh | 56 | 0.992 |
| Maharashtra | 120 | 0.989 |
| Tamilnadu | 70 | 0.986 |
| Jammu Kashmir | 189 | 0.985 |
| Uttar Pradesh | 2 | 0.983 |
| Uttarakhand | 66 | 0.982 |
| Odisha | 29 | 0.978 |
| Rajasthan | 3 | 0.976 |
| Puducherry | 35 | 0.974 |
| Madhya Pradesh | 237 | 0.971 |
| West Bengal | 171 | 0.971 |
| Chhattisgarh | 92 | 0.961 |
| Jharkhand | 22 | 0.956 |
| Himachal Pradesh | 48 | 0.948 |
| Assam | 2 | 0.933 |
| Tripura | 5 | 0.915 |
| Karnataka | 131 | 0.895 |
| Sikkim | 8 | 0.863 |
| Punjab | 692 | 0.861 |
| Goa | 5 | 0.8 |
| Meghalaya | 15 | 0.798 |

Irrespective of the high frequency of this variant in Punjab, we do not see a high diversity of this variant, similar to Chandigarh and New Delhi (Table 1). This suggests that the spread of the virus in Punjab was rapid and associated with a limited number of founder events. On the other hand, farmers’ protests in Punjab and other regions caused an exponential increase in frequency in Punjab, with the majority of the farmers hailing from Punjab and Haryana. Rallies were staged in several parts of north India, highlighting the critical role of social dynamics in the SARS-CoV-2 pandemic and the rise of the second wave in India. In April, cases of the Delta variant were more prevalent than those of the alpha type, and the frequency percentage in most locations declined. Concerning the alpha variant, there existed a strong link between Delhi and Punjab. The outbreak in Delhi in April 2021 was preceded by outbreaks in Maharashtra, Kerala, and Punjab, and we can say that the outbreak in Delhi in 2021 was fueled by alpha alone (*GISAID - Initiative*, 2008).

Thus, this study outlines the need for phylogenetics to understand a virus variant’s spatial and temporal distribution. Virus variant alpha was predominant in north India compared to the southern part. The rapid expansion of this variant has had limited founders in Punjab and was likely to be associated with the farmers’ agitation. Its moderate presence in the south was likely due to a more virulent strain delta which has eventually taken the complete landscape of India after that.

The outcome of this study is dependent on the number of sequences and available samples. However, there are places where proper testing, collecting, sequencing and submission of samples have not been adequately made, which may impact the outcome.

## Data Availability

All datasets generated for this study are included in the article/Supplementary Material

https://drive.google.com/drive/folders/1ld7ghGsxDWWYPHQokotrFeNL_t4YnOeh?usp=sharing

## Acknowledgements

This work is supported by ICMR ad-hoc grant (2021-6389). GC and VNM is supported by ICMR (2021-11289) and Faculty IOE grant BHU (6031). PPS is supported by CSIR SRF fellowship. RKP is supported by ICMR-SRF fellowship. AS is supported by UGC-CAS fellowship. RKM is supported by Mahamana post doctoral fellowship BHU.

## Author contributions

GC conceived and designed this study. JP, RKP, YP, PPS, AS, RM, BK, NR, VNM, PS, PrS collected the data. JP, RKP, YP, PPS analyzed the data. JP, PPS and GC wrote the manuscript from the inputs of other co-authors. All authors contributed to the article and approved the submitted version.

## Data Availability Statement

All datasets generated for this study are included in the article/Supplementary Material.

## Competing interests

The authors declare no competing interests.

**Supplementary Fig. 1** The Maximum Likelihood (ML) tree showing the diverse placement of three major regions of alpha variant in India.

**Supplementary Fig. 2**. Frequency plot of the regions affected by alpha variant from Dec 2020 to May 2021.

Supplementary Table 1. The details of samples studied and the frequency of alpha variant in various regions of India.

## References

A. Sarah Walker, O.G., Emma Pritchard, Joel Jones, & Thomas House. (2021). Tracking the Emergence of SARS-CoV-2 Alpha Variant in the United Kingdom. NEW ENGLAND JOURNAL of MEDICINE. https://doi.org/10.1056/NEJMc2103227

Anshika Srivastava, A. B., Debashurti Das, Rudra Kumar, Vanya Singh, Nargis Khanam, Nikhil Srivastava, Prajjval Pratap Singh, Pavan Kumar Dubey, Abhishek Pathak, Pranav Gupta, Niraj Rai, & Gyaneshwer Chaubey. (2020). Genetic Association of ACE2 rs2285666 Polymorphism With COVID-19 Spatial Distribution in India. Frontiers in Genetics. https://doi.org/10.3389/fgene.2020.564741

Ewen Callaway. (2021). Delta coronavirus variant: Scientists brace for impact. Nature. https://doi.org/10.1038/d41586-021-01696-3

Finlay Campbell, B. A., Henry Laurenson-Schafer, Yuka Jinnai, Franck Konings, Neale Batra, Boris Pavlin, Katelijn Vandemaele, Maria D Van Kerkhove, Thibaut Jombart, Oliver Morgan, & Olivier le Polain de Waroux. (2021). Increased transmissibility and global spread of SARS-CoV-2 variants of concern. PubMed. https://doi.org/10.2807/1560-7917.ES.2021.26.24.2100509

Fu, Yun-Xin, and Wen-Hsiung Li 1993 Statistical Tests of Neutrality of Mutations. Genetics 133(3). Oxford University Press: 693–709.

GISAID - Initiative. (2008). GISAID. https://www.gisaid.org/

Gobeil, K. J., Shana McDowell, Katayoun Mansouri, Robert Parks, Victoria Stalls, Megan F Kopp, Kartik Manne, Kevin Saunders, Robert J Edwards, Barton F Haynes, Rory C Henderson, & Priyamvada Acharya. (2021). Effect of natural mutations of SARS-CoV-2 on spike structure, conformation, and antigenicity. PubMed. https://doi.org/10.1101/2021.03.11.435037

Google sheets. (2009). Google. https://www.google.com/sheets/about/

Julio Rozas, A. F.-M., DelBarrio, Guirao-Rico, & Pablo Librado. (2019). DnaSP (6.12.03) [Computer software]. Universitat de Barcelona. http://www.ub.edu/dnasp/

Lucy, M. B., Ann-Kathrin Reuschl, Lorena Zuliani, & Nevan J Krogan. (2021). Evolution of enhanced innate immune evasion by the SARS-CoV-2 B.1.1.7 UK variant. PubMed. https://doi.org/10.1101/2021.06.06.446826

MAFFT (Version 7). (2020). [Computer software]. Multiple alignment program for amino acid or nucleotide sequences

MEGAX (10.2.4). (2021). [Computer software]. Pennsylvania State University. https://www.megasoftware.net/

Mirko Lorenz, Nicolas Kayser-Bril, & Gregor Aisch. (2012). Datawrapper. https://www.datawrapper.de/ MOHFW-GoI. (n.d.). mohfw.gov.in

Qianqian Zhang, F. Y., Xiang, R., Shanshan Huo, Yunjiao Zhou, & Shibo Jiang. (2021). Molecular mechanism of interaction between SARS-CoV-2 and host cells and interventional therapy. Nature. https://www.nature.com/articles/s41392-021-00653-w

Robert Challen, E. B.-P., Jonathan M Read, Louise Dyson, Krasimira Tsaneva-Atanasova, & Leon Danon. (2021). Risk of mortality in patients infected with SARS-CoV-2 variant of concern 202012/1: Matched cohort study. The BMJ. https://doi.org/10.1136/bmj.n579

Rossana C. Jaspe, C.L.L., YoneiraSulbaran, Zoila C. Moros, PierinaD’Angelo LieskaRodríguez, José LuisZambrano, MarianaHidalgo, EsmeraldaVizzi VíctorAlarcón, MarwanAguilar, & Domingo J. Garzaro. (2021). Introduction and rapid dissemination of SARS-CoV-2 Gamma Variant of Concern in Venezuela. ScienceDirect, 96. https://doi.org/10.1016/j.meegid.2021.105147

WHO. (n.d.). Tracking SARS-CoV-2 variants. https://www.who.int/en/activities/tracking-SARS-CoV-2-variants/

Singh, Keshav K, Gyaneshwer Chaubey, Jake Y Chen, and Prashanth Suravajhala 2020 Decoding SARS-CoV-2 Hijacking of Host Mitochondria in COVID-19 Pathogenesis. American Journal of Physiology-Cell Physiology 319(2). American Physiological Society Bethesda, MD: C258–C267.

William T. Harvey, D.L.R., Alessandro M. Carabelli, Ben Jackson, Ravindra K. Gupta, Emma C. Thomson, Ewan M. Harrison, Catherine Ludden, Richard Reeve, Andrew Rambaut, & Sharon J. Peacock. (2021). SARS-CoV-2 variants, spike mutations and immune escape. Nature Reviews Microbiology. https://www.nature.com/articles/s41579-021-00573-0

Wu, J. T., Leung, K., Bushman, M., Kishore, N., Niehus, R., de Salazar, P. M., Cowling, B. J., Lipsitch, M., & Leung, G. M. (2020). Estimating clinical severity of COVID-19 from the transmission dynamics in Wuhan, China. Nature Medicine, 1–5.

